# Trends in Cardiometabolic Disease and Health-Related Quality of Life in the United States, 2001-2022

**DOI:** 10.64898/2026.02.20.26346754

**Authors:** Danwei Yang, David D. Kim

## Abstract

**Objectives:** To examine associations between cardiometabolic conditions and health-related quality of life (HRQoL) and to evaluate whether condition-associated HRQoL changed from 2001 to 2022.

**Methods:** We analyzed nationally representative data from U.S. adults aged ≥18 years in the Medical Expenditure Panel Survey, 2001-2022. Survey years without BMI data (2017, 2019, 2021) were excluded. EQ-5D utilities were mapped from SF-12 scores using a validated algorithm. For each survey year, survey-weighted multivariable regression models estimated associations of sociodemographic characteristics, BMI, and cardiometabolic conditions (diabetes, heart disease, high blood pressure, high cholesterol, obesity, stroke) with HRQoL measured by EQ-5D. Temporal changes in condition-associated HRQoL decrements were assessed using meta-regression across years. Associations in recent survey years were summarized using pooled estimates from 2015, 2016, 2018, and 2022.

**Results:** Overall HRQoL improved from 2001 to 2022 across age groups, with the largest improvement among older adults. In pooled analyses, stroke was associated with the largest adjusted HRQoL decrement (−0.0714), followed by heart disease (−0.0503), diabetes (−0.0427), high blood pressure (−0.0328), obesity (−0.0305), and high cholesterol (−0.0236). Additional adjustment for BMI attenuated condition-associated decrements, most notably for obesity (−0.0305 to −0.0183), diabetes (−0.0427 to −0.0414), and high blood pressure (−0.0328 to −0.0316). Over time, diabetes- and heart disease-associated decrements attenuated linearly (diabetes: - 0.0489 in 2001 to −0.0406 in 2022; heart disease: −0.0591 to −0.0493). High blood pressure (−0.0337 in 2001, −0.0415 in 2012, −0.0306 in 2022) and obesity (−0.0305 in 2001, −0.0283 in 2012, −0.0367 in 2022) showed nonlinear patterns.

**Conclusions:** Condition-associated HRQoL decrements varied over time, and recent-year utility estimates are recommended for population health research. HRQoL decrements for diabetes and heart disease attenuated, consistent with improvements in treatment and survival. High blood pressure-associated were lowest around 2012, and obesity-associated became more negative after 2012, consistent with worsening blood pressure control and obesity severity.

## 1. Introduction

Health-related quality of life (HRQoL) reflects how health influences daily functioning, psychological well-being, social participation, and perceived overall health.^1,2^ The EuroQol 5-Dimension (EQ-5D) instrument describes health across five dimensions and converts responses into a single preference-based utility index scaled from 0 (death) to 1 (full health).^3^ EQ-5D utilities are commonly used in population health research and economic evaluation by combining utility with survival to estimate quality-adjusted life years (QALYs).^4,5^

Cardiometabolic conditions, such as cardiovascular disease, type 2 diabetes, and obesity, share common risk factors and contribute substantially to morbidity and mortality in the U.S., with this burden expected to persist.^6–8^ These conditions are associated with limitations in physical functioning and daily activities and with lower HRQoL.^9–11^ Over the past two decades, advances in prevention and treatment may have improved symptoms and functioning among adults living with cardiometabolic conditions.^12–15^ At the same time, improved survival has increased the number of people living with chronic complications and multimorbidity, alongside rising obesity prevalence.^16^ Together, these forces make population-level trends in HRQoL uncertain.^17^

Despite extensive research on HRQoL associated with cardiometabolic conditions, gaps remain in understanding population-level changes over time. First, much of the evidence comes from clinical cohorts using condition-specific patient-reported outcome measures (eg, the Kansas City Cardiomyopathy Questionnaire for heart failure), which are valuable for within-condition assessment but are not designed for cross-condition comparisons or population level estimation.^18–22^ Second, although nationally representative utility data (eg, from the Medical Expenditure Panel Survey [MEPS]) support population-level estimation across cardiometabolic conditions, many published estimates rely on earlier periods and may not reflect more recent patterns of prevention, treatment, and survival.^23,24^ Third, because cardiometabolic conditions share common risk factors, inconsistent adjustment for obesity or body mass index (BMI) can confound condition-specific HRQoL estimates and limit comparability across conditions.^25^

To assess whether HRQoL associated with cardiometabolic conditions has changed amid evolving prevention and treatment in the United States, we used nationally representative data to (1) estimate associations of sociodemographic characteristics, BMI, and cardiometabolic conditions (diabetes, heart disease, high blood pressure, high cholesterol, stroke, and obesity) with HRQoL, measured by EQ-5D utility; (2) evaluate temporal trends in condition-associated HRQoL decrements; and (3) provide updated nationally representative HRQoL utility estimates for use in population health research.

## 2. Methods

### 2.1 Data

We used the MEPS, a nationally representative survey of the U.S. civilian noninstitutionalized population that has collected the 12-Item Short Form Survey (SF-12) since 2001, EQ-5D measurement from 2000 to 2003, and detailed sociodemographic characteristics and chronic condition indicators.^26^ We analyzed adults aged ≥18 years in the 2001-2022 MEPS Full-Year Consolidated Public Use Files. BMI and HRQoL measures are obtained from the Self-Administered Questionnaire (SAQ). Analyses were limited to SAQ respondents and used SAQ person-level survey weights. Survey years 2017, 2019, and 2021, which did not collect BMI information, were excluded. To avoid conflating secular trends with pandemic-related changes, we excluded 2020 from primary analyses and evaluated it in sensitivity analyses.

### 2.2. Cardiometabolic Conditions

MEPS priority condition indicators were used to identify diagnosed diabetes, heart disease, high blood pressure, high cholesterol (available starting in 2005), and stroke. BMI categories were defined using self-reported BMI with standard cut points: underweight (<18.5 kg/m²), normal weight (18.5-24.9), overweight (25.0-29.9), obesity class I (30.0-34.9), obesity class II (35.0-39.9), and obesity class III (≥40).^27^

### 2.3. Mapping EQ-5D From SF-12

To obtain EQ-5D utilities consistently across study years, we applied a validated mapping algorithm that predicts EQ-5D utilities from the SF-12.^28^ We applied a censored least absolute deviations (CLAD) specification algorithm, which accommodates the bounded distribution and ceiling effects of utility measures and is robust to non-normality and heteroskedasticity. The mapping model included the Physical Component Summary (PCS-12), Mental Component Summary (MCS-12), sociodemographic characteristics (age, sex, race/ethnicity, income, education), and the number of chronic conditions (NCC) based on MEPS priority conditions. Predicted EQ-5D values were bounded to keep utilities within the conventional 0-1 scale.

### 2.4 Analysis

We used survey-weighted multivariable linear regression models to estimate associations of sociodemographic characteristics, cardiometabolic conditions, and BMI with HRQoL, measured using the EQ-5D index. Linear models were selected to estimate adjusted mean differences in HRQoL utilities across population subgroups and over time, yielding directly interpretable coefficients for condition-specific HRQoL decrements and temporal comparisons. Because the association between BMI and HRQoL is nonlinear, BMI was modeled as a continuous variable centered at 25 kg/m², with a quadratic term to capture nonlinearity and to facilitate interpretation at a clinically meaningful reference point.^29^

We estimated four nested specifications: (1) sociodemographic covariates only (age group, sex, race/ethnicity, education, income); (2) sociodemographic covariates plus cardiometabolic conditions (diabetes, heart disease, high blood pressure, high cholesterol, obesity, stroke); (3) sociodemographic covariates + BMI (centered at 25 kg/m²) + BMI²; and (4) sociodemographic covariates plus cardiometabolic conditions, BMI, and BMI². Comparing coefficients and model fit across these nested specifications allowed us to quantify overlapping versus independent associations of BMI and cardiometabolic conditions with HRQoL and to evaluate whether observed sociodemographic gradients persisted after adjustment.

We evaluated four prespecified model specifications for each survey year and estimated pooled associations using selected recent survey years (2015, 2016, 2018, and 2022) to improve precision and reflect more recent patterns. We fit year-specific models to estimate condition-specific HRQoL decrements, defined as the adjusted difference in HRQoL between adults with versus without each cardiometabolic condition (ie, the regression coefficient for the condition indicator). We then used meta-regression with calendar year as the moderator to test temporal trends in these decrements, evaluating linear trends and adding quadratic terms when indicated. Coefficients moving toward 0 were interpreted as attenuation (smaller decrements), whereas coefficients moving away from 0 were interpreted as worsening (larger decrements).

## 3. Results

### 3.1 Population Characteristics and HRQoL, 2001-2022

From 2001 to 2022, HRQoL improved overall, with a temporary decline in 2020 (Figure 1). Mean EQ-5D was 0.877 (95% CI, 0.874−0.880) in 2001-2012, increased to 0.888 (95% CI, 0.885−0.891) in 2018, then declined to 0.882 (95% CI, 0.880−0.886) in 2020; however, the 2020 estimate remained higher than the 2001-2012 average. MCS-12 demonstrated a similar pattern, rising from average 51.004 (95% CI, 50.815-51.194) in 2001-2012 to 52.261 (95% CI, 52.064-52.458) in 2018, and declining to 51.356 (95% CI, 51.113-51.599) in 2020. PCS-12 increased over time and showed little change during 2020, rising from 49.114 (95% CI, 48.917-49.311) in 2001 to 50.155 (95% CI, 49.853-50.457) in 2022.

**Figure 1.**
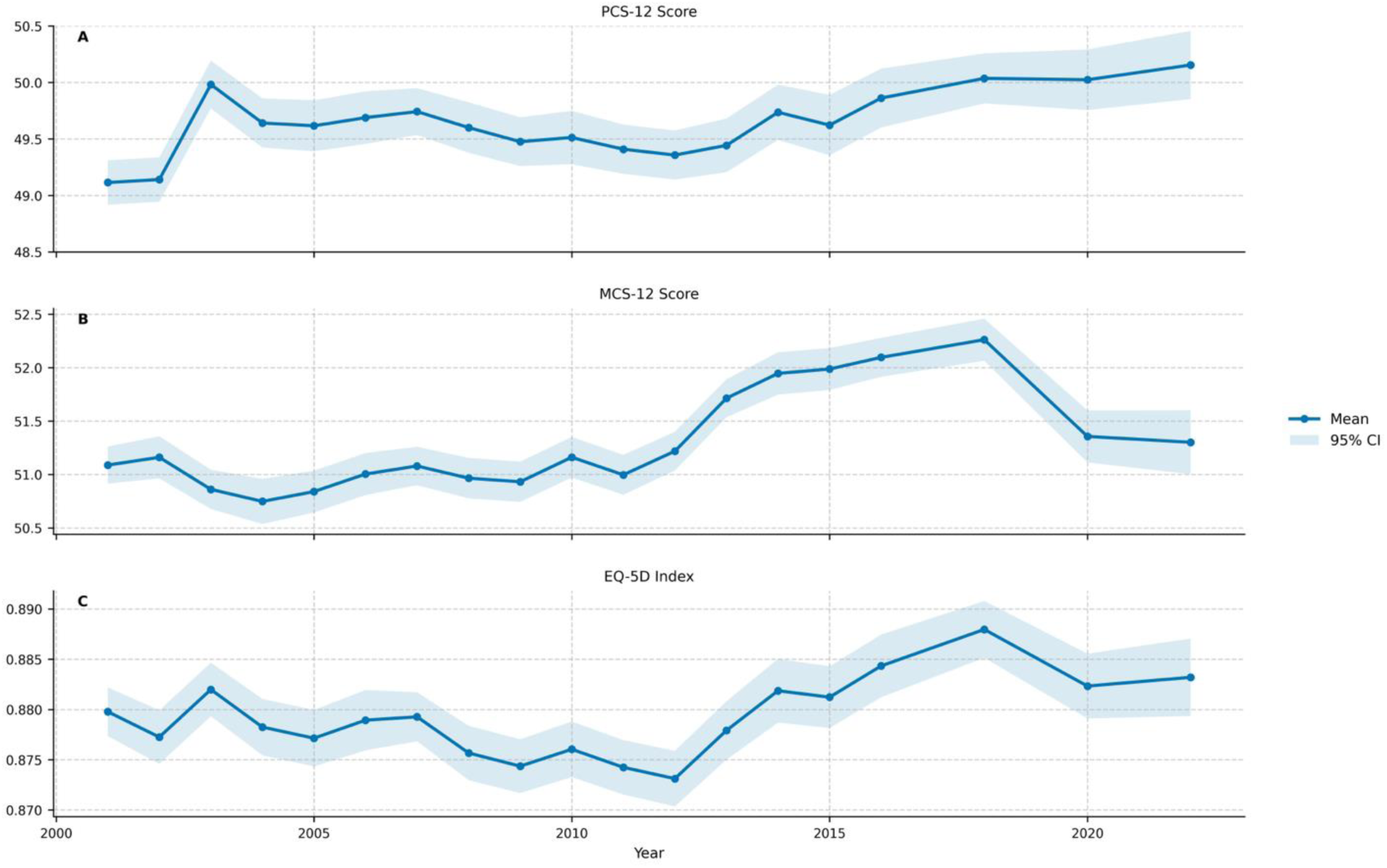
Trends in PCS-12, MCS-12, and EQ-5D index among U.S. adults, MEPS 2001-2022.

HRQoL differed by sociodemographic characteristics, cardiometabolic conditions, and BMI category (Table 1), with survey-weighted estimates from 2001-2022 representing 229.3 million U.S. adults. Mean HRQoL was higher among younger than older adults (0.915 for ages 30-39 vs 0.840 for ages 60-69) and among men than women (0.891 vs 0.869). By race and ethnicity, mean HRQoL was highest among non-Hispanic Asian adults (0.910), followed by Hispanic (0.895), non-Hispanic White (0.876), and non-Hispanic Black adults (0.870). HRQoL increased with socioeconomic status, from 0.819 among adults at ≤ 100% federal poverty level to 0.917 among adults at ≥ 400%, and from 0.841 among adults with no degree to 0.917 among adults with Masters or Doctorates. Across cardiometabolic conditions, mean HRQoL was lowest among adults with stroke (0.718), followed by diabetes (0.769), heart disease (0.778), high blood pressure (0.810), high cholesterol (0.822), and obesity (0.842). HRQoL exhibited an inverse U-shaped pattern across BMI categories: adults in the normal-weight category had the highest mean utility (0.904), whereas underweight (0.862), overweight (0.890), and obesity (≤0.858) had lower utilities, with progressively lower values at higher obesity classes.

**Table 1.** Weighted sociodemographic characteristics, comorbidity burden, and HRQoL measures among U.S. adults, MEPS 2001-2022 (N=229,317,502)

|  | % of<br>population | Age (Years) |  | NCC |  | PCS |  | MCS |  | EQ-5D |  |
| --- | --- | --- | --- | --- | --- | --- | --- | --- | --- | --- | --- |
|  |  | Mean | SE | Mean | SE | Mean | SE | Mean | SE | Mean | SE |
| MEPS Weighted Population | 100.00% | 46.87 | 0.11 | 1.86 | 0.01 | 49.651 | 0.048 | 51.320 | 0.035 | 0.879 | 0.001 |
| Age group |  |  |  |  |  |  |  |  |  |  |  |
| 18-29 | 20.88% | 23.71 | 0.03 | 0.64 | 0.01 | 54.374 | 0.041 | 51.333 | 0.063 | 0.932 | 0.001 |
| 30-39 | 17.74% | 34.49 | 0.02 | 1.03 | 0.01 | 53.109 | 0.052 | 50.832 | 0.060 | 0.915 | 0.001 |
| 40-49 | 18.02% | 44.50 | 0.02 | 1.55 | 0.01 | 51.008 | 0.064 | 50.780 | 0.057 | 0.891 | 0.001 |
| 50-59 | 17.56% | 54.36 | 0.02 | 2.30 | 0.02 | 48.372 | 0.082 | 51.059 | 0.062 | 0.862 | 0.001 |
| 60-69 | 13.12% | 64.15 | 0.02 | 3.07 | 0.02 | 45.778 | 0.106 | 52.218 | 0.072 | 0.840 | 0.001 |
| 70-79 | 8.07% | 74.07 | 0.02 | 3.61 | 0.02 | 42.624 | 0.117 | 52.703 | 0.078 | 0.810 | 0.001 |
| 80+ | 4.62% | 83.26 | 0.02 | 3.67 | 0.02 | 37.875 | 0.138 | 51.264 | 0.117 | 0.756 | 0.002 |
| Sex |  |  |  |  |  |  |  |  |  |  |  |
| Female | 51.74% | 47.59 | 0.11 | 1.93 | 0.01 | 48.953 | 0.057 | 50.476 | 0.041 | 0.869 | 0.001 |
| Male | 48.26% | 46.09 | 0.11 | 1.79 | 0.01 | 50.399 | 0.049 | 52.224 | 0.038 | 0.891 | 0.001 |
| Race |  |  |  |  |  |  |  |  |  |  |  |
| Non-Hispanic White | 66.74% | 48.92 | 0.13 | 2.01 | 0.01 | 49.416 | 0.058 | 51.229 | 0.042 | 0.876 | 0.001 |
| Non-Hispanic Black | 11.49% | 44.29 | 0.16 | 1.95 | 0.02 | 48.867 | 0.093 | 51.419 | 0.074 | 0.870 | 0.001 |
| Hispanic | 14.42% | 40.88 | 0.15 | 1.36 | 0.01 | 50.929 | 0.080 | 51.485 | 0.072 | 0.895 | 0.001 |
| Non-Hispanic Asian/Pacific Islander | 5.23% | 44.33 | 0.39 | 1.09 | 0.03 | 51.490 | 0.127 | 52.360 | 0.111 | 0.910 | 0.002 |
| Non-Hispanic other | 2.12% | 43.29 | 0.32 | 2.17 | 0.05 | 48.059 | 0.248 | 49.944 | 0.204 | 0.855 | 0.003 |
| Education |  |  |  |  |  |  |  |  |  |  |  |
| No degree | 14.05% | 46.02 | 0.18 | 1.94 | 0.02 | 46.917 | 0.102 | 49.937 | 0.075 | 0.841 | 0.001 |
| High school diploma | 48.23% | 46.44 | 0.12 | 1.95 | 0.01 | 48.999 | 0.053 | 51.224 | 0.043 | 0.871 | 0.001 |
| Other degree | 6.83% | 47.02 | 0.20 | 1.93 | 0.02 | 50.061 | 0.102 | 51.371 | 0.092 | 0.885 | 0.001 |
| Bachelor's degree | 17.87% | 46.31 | 0.17 | 1.58 | 0.01 | 52.156 | 0.063 | 51.983 | 0.059 | 0.912 | 0.001 |
| Masters or Doctorate degree | 9.77% | 50.96 | 0.19 | 1.75 | 0.02 | 52.203 | 0.075 | 52.595 | 0.071 | 0.917 | 0.001 |
| Income (% poverty level) |  |  |  |  |  |  |  |  |  |  |  |
| Poor (<100%) | 11.18% | 43.56 | 0.17 | 2.04 | 0.02 | 46.182 | 0.094 | 47.861 | 0.088 | 0.819 | 0.001 |
| Near poor (100%-124%) | 4.01% | 48.32 | 0.29 | 2.22 | 0.03 | 46.005 | 0.144 | 49.162 | 0.115 | 0.829 | 0.002 |
| Low income (125%-199%) | 12.90% | 47.64 | 0.19 | 2.04 | 0.02 | 47.477 | 0.084 | 50.210 | 0.072 | 0.852 | 0.001 |
| Middle income (200%-399%) | 30.22% | 45.83 | 0.13 | 1.82 | 0.01 | 49.807 | 0.057 | 51.441 | 0.049 | 0.882 | 0.001 |
| High income (>=400%) | 41.69% | 48.14 | 0.12 | 1.76 | 0.01 | 51.491 | 0.047 | 52.710 | 0.040 | 0.907 | 0.001 |
| Condition |  |  |  |  |  |  |  |  |  |  |  |
| Diabetes | 9.14% | 60.99 | 0.14 | 4.65 | 0.02 | 40.713 | 0.111 | 49.491 | 0.093 | 0.769 | 0.001 |
| Heart Disease | 13.05% | 61.81 | 0.19 | 4.35 | 0.02 | 41.402 | 0.111 | 49.668 | 0.085 | 0.778 | 0.001 |
| High Blood Pressure | 30.70% | 59.48 | 0.12 | 3.74 | 0.01 | 43.999 | 0.072 | 50.510 | 0.056 | 0.810 | 0.001 |
| High Cholesterol | 24.57% | 59.26 | 0.13 | 3.83 | 0.02 | 45.121 | 0.093 | 50.736 | 0.062 | 0.822 | 0.001 |
| Obesity | 28.66% | 48.00 | 0.11 | 3.12 | 0.01 | 46.801 | 0.069 | 50.519 | 0.051 | 0.842 | 0.001 |
| Stroke | 3.36% | 66.88 | 0.20 | 5.10 | 0.03 | 36.790 | 0.157 | 47.787 | 0.160 | 0.718 | 0.002 |
| BMI (kg/m<sup>2</sup>) |  |  |  |  |  |  |  |  |  |  |  |
| underweight (<18.5) | 4.71% | 46.52 | 0.26 | 1.37 | 0.02 | 48.092 | 0.137 | 50.446 | 0.114 | 0.862 | 0.002 |
| normal weight (18.5-24.9) | 33.17% | 44.37 | 0.14 | 1.15 | 0.01 | 51.707 | 0.056 | 51.548 | 0.045 | 0.904 | 0.001 |
| overweight (25.0-29.9) | 33.46% | 48.43 | 0.13 | 1.57 | 0.01 | 50.272 | 0.052 | 51.902 | 0.045 | 0.890 | 0.001 |
| obesity class I (30.0-34.9) | 17.14% | 48.65 | 0.13 | 2.96 | 0.01 | 48.099 | 0.070 | 51.088 | 0.059 | 0.858 | 0.001 |
| obesity class II (35.0-39.9) | 7.05% | 47.72 | 0.16 | 3.26 | 0.02 | 46.066 | 0.108 | 50.295 | 0.089 | 0.833 | 0.001 |
| obesity class III (≥40) | 4.48% | 45.97 | 0.17 | 3.54 | 0.03 | 42.990 | 0.145 | 48.696 | 0.127 | 0.795 | 0.002 |

Mean HRQoL values increased over time across age groups, with larger improvements among older adults (particularly ages 70-79 and ≥80) (Figure 2). HRQoL also improved over time across sex, race and ethnicity, education, and income subgroups over cardiometabolic conditions, and trajectories were broadly similar across subgroups (eg, men and women showed comparable trends). Differences between age groups became approximately parallel by 2015. Thus, we pooled 2015, 2016, 2018, and 2022 to estimate associations in later study years with improved precision.

**Figure 2.**
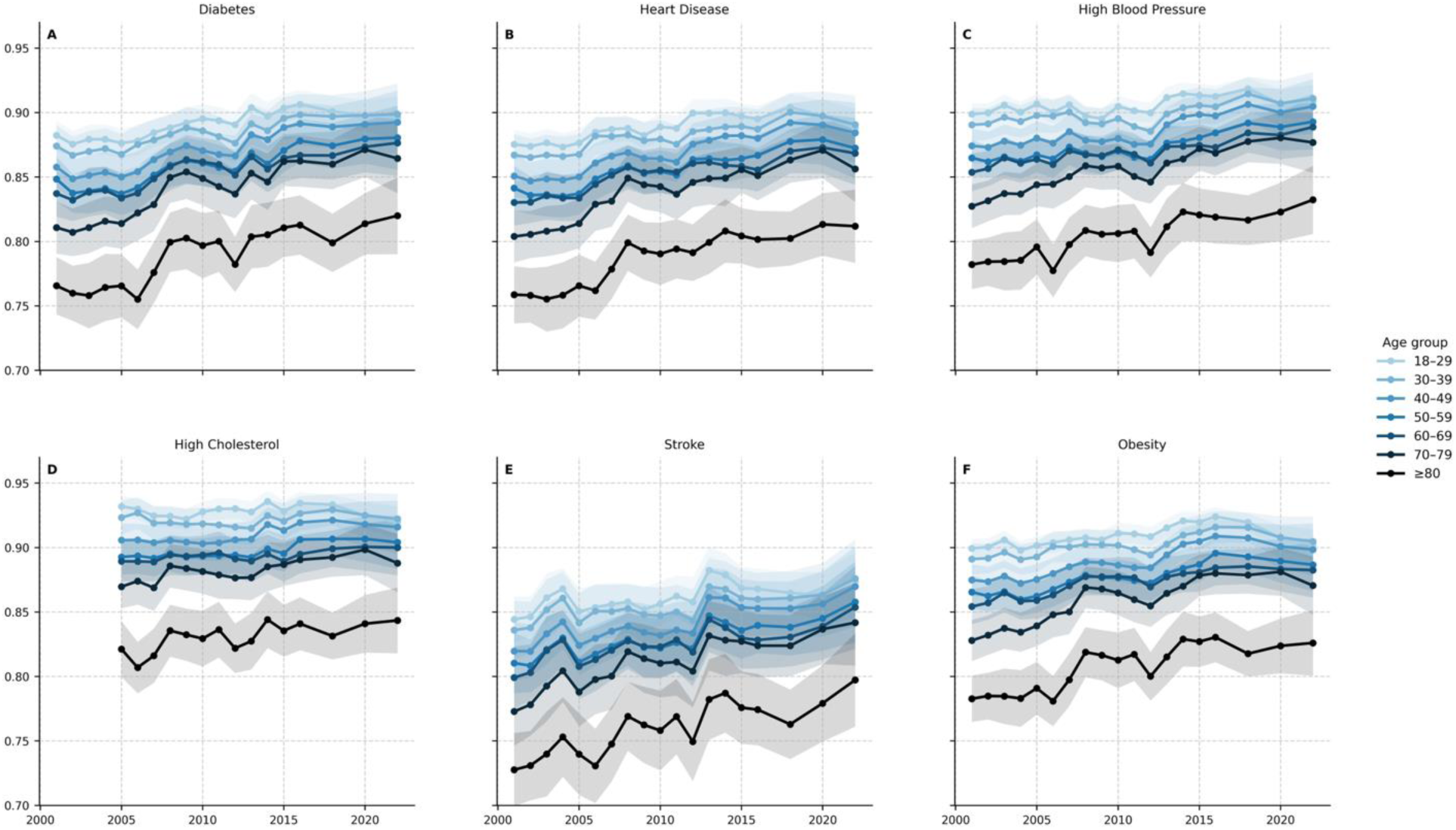
Predicted mean HRQoL by age group across cardiometabolic conditions, 2001-2022 (Model 2)

### 3.2 Association between Sociodemographic, Cardiometabolic Conditions, BMI and HRQoL

Compared with the sociodemographic-only specification (Model 1), adding cardiometabolic conditions (Model 2), BMI with a quadratic term (Model 3), or both (Model 4) improved model fit in the aggregated data for 2015, 2016, 2018, and 2022 (Table 2) and across all years (Supplementary Tables A-S). Although inclusion of BMI yielded only modest incremental improvement in model fit, the BMI terms were statistically significant in Model 3 and 4, supporting an independent association between BMI and HRQoL after adjustment for cardiometabolic conditions.

**Table 2.** Survey-weighted multivariable linear regression of HRQoL on sociodemographic characteristics, BMI, and cardiometabolic conditions, MEPS 2015, 2016.

|  | Model 1 |  |  | Model 2 |  |  | Model 3 |  |  | Model 4 |  |  |
| --- | --- | --- | --- | --- | --- | --- | --- | --- | --- | --- | --- | --- |
|  | Beta | (SE) | *p* | Beta | (SE) | *p* | Beta | (SE) | *p* | Beta | (SE) | *p* |
| Intercept | 0.8493 | 0.0027 | <0.0001 | 0.8684 | 0.0025 | <0.0001 | 0.8536 | 0.0030 | <0.0001 | 0.8683 | 0.0025 | <0.0001 |
| Age group |  |  |  |  |  |  |  |  |  |  |  |  |
| 18-29 |  |  |  |  |  |  |  |  |  |  |  |  |
| 30-39 | -0.0264 | 0.0018 | <0.0001 | -0.0146 | 0.0016 | <0.0001 | -0.0214 | 0.0020 | <0.0001 | -0.0140 | 0.0016 | <0.0001 |
| 40-49 | -0.0484 | 0.0019 | <0.0001 | -0.0240 | 0.0017 | <0.0001 | -0.0415 | 0.0022 | <0.0001 | -0.0234 | 0.0018 | <0.0001 |
| 50-59 | -0.0764 | 0.0019 | <0.0001 | -0.0375 | 0.0017 | <0.0001 | -0.0701 | 0.0022 | <0.0001 | -0.0373 | 0.0017 | <0.0001 |
| 60-69 | -0.0994 | 0.0023 | <0.0001 | -0.0434 | 0.0021 | <0.0001 | -0.0943 | 0.0025 | <0.0001 | -0.0438 | 0.0021 | <0.0001 |
| 70-79 | -0.1197 | 0.0027 | <0.0001 | -0.0482 | 0.0025 | <0.0001 | -0.1165 | 0.0028 | <0.0001 | -0.0492 | 0.0024 | <0.0001 |
| 80+ | -0.1653 | 0.0032 | <0.0001 | -0.0900 | 0.0032 | <0.0001 | -0.1676 | 0.0031 | <0.0001 | -0.0917 | 0.0033 | <0.0001 |
| Sex |  |  |  |  |  |  |  |  |  |  |  |  |
| Female |  |  |  |  |  |  |  |  |  |  |  |  |
| Male | 0.0131 | 0.0009 | <0.0001 | 0.0173 | 0.0009 | <0.0001 | 0.0127 | 0.0009 | <0.0001 | 0.0171 | 0.0009 | <0.0001 |
| Race |  |  |  |  |  |  |  |  |  |  |  |  |
| Non-Hispanic White |  |  |  |  |  |  |  |  |  |  |  |  |
| Non-Hispanic Black | 0.0081 | 0.0019 | <0.0001 | 0.0139 | 0.0017 | <0.0001 | 0.0124 | 0.0020 | <0.0001 | 0.0145 | 0.0017 | <0.0001 |
| Hispanic | 0.0292 | 0.0018 | <0.0001 | 0.0253 | 0.0016 | <0.0001 | 0.0292 | 0.0018 | <0.0001 | 0.0254 | 0.0016 | <0.0001 |
| Non-Hispanic Asian/Pacific Islander | 0.0189 | 0.0026 | <0.0001 | 0.0090 | 0.0025 | 0.0003 | 0.0107 | 0.0028 | 0.0001 | 0.0080 | 0.0025 | 0.0014 |
| Non-Hispanic other | -0.0140 | 0.0036 | 0.0001 | -0.0077 | 0.0033 | 0.0216 | -0.0118 | 0.0035 | 0.0009 | -0.0076 | 0.0033 | 0.0213 |
| Education |  |  |  |  |  |  |  |  |  |  |  |  |
| No degree |  |  |  |  |  |  |  |  |  |  |  |  |
| High school diploma | 0.0132 | 0.0019 | <0.0001 | 0.0135 | 0.0017 | <0.0001 | 0.0158 | 0.0018 | <0.0001 | 0.0140 | 0.0017 | <0.0001 |
| Other degree | 0.0201 | 0.0028 | <0.0001 | 0.0198 | 0.0026 | <0.0001 | 0.0226 | 0.0028 | <0.0001 | 0.0203 | 0.0026 | <0.0001 |
| Bachelor's degree | 0.0410 | 0.0021 | <0.0001 | 0.0335 | 0.0019 | <0.0001 | 0.0410 | 0.0021 | <0.0001 | 0.0337 | 0.0019 | <0.0001 |
| Masters or Doctorate degree | 0.0531 | 0.0024 | <0.0001 | 0.0436 | 0.0022 | <0.0001 | 0.0518 | 0.0025 | <0.0001 | 0.0436 | 0.0022 | <0.0001 |
| Income (% poverty level) |  |  |  |  |  |  |  |  |  |  |  |  |
| Poor (<100%) |  |  |  |  |  |  |  |  |  |  |  |  |
| Near poor (100%-124%) | 0.0224 | 0.0037 | <0.0001 | 0.0211 | 0.0033 | <0.0001 | 0.0226 | 0.0036 | <0.0001 | 0.0212 | 0.0033 | <0.0001 |
| Low income (125%-199%) | 0.0366 | 0.0027 | <0.0001 | 0.0344 | 0.0024 | <0.0001 | 0.0372 | 0.0025 | <0.0001 | 0.0344 | 0.0024 | <0.0001 |
| Middle income (200%-399%) | 0.0602 | 0.0023 | <0.0001 | 0.0539 | 0.0021 | <0.0001 | 0.0604 | 0.0023 | <0.0001 | 0.0538 | 0.0021 | <0.0001 |
| High income (>=400%) | 0.0819 | 0.0023 | <0.0001 | 0.0707 | 0.0021 | <0.0001 | 0.0800 | 0.0023 | <0.0001 | 0.0702 | 0.0021 | <0.0001 |
| Condition |  |  |  |  |  |  |  |  |  |  |  |  |
| Diabetes |  |  |  | -0.0427 | 0.0022 | <0.0001 |  |  |  | -0.0414 | 0.0023 | <0.0001 |
| Heart Disease |  |  |  | -0.0503 | 0.0019 | <0.0001 |  |  |  | -0.0502 | 0.0019 | <0.0001 |
| High Blood Pressure |  |  |  | -0.0328 | 0.0015 | <0.0001 |  |  |  | -0.0316 | 0.0015 | <0.0001 |
| High Cholesterol |  |  |  | -0.0236 | 0.0013 | <0.0001 |  |  |  | -0.0234 | 0.0013 | <0.0001 |
| Obesity |  |  |  | -0.0305 | 0.0012 | <0.0001 |  |  |  | -0.0183 | 0.0038 | <0.0001 |
| Stroke |  |  |  | -0.0714 | 0.0036 | <0.0001 |  |  |  | -0.0716 | 0.0036 | <0.0001 |
| BMI |  |  |  |  |  |  | -0.0022 | 0.0003 | <0.0001 | -0.0009 | 0.0003 | 0.0011 |
| BMI*BMI |  |  |  |  |  |  | 0.0000 | 0.0000 | 0.0001 | 0.0000 | 0.0000 | 0.0030 |

Adjustment for cardiometabolic conditions substantially attenuated the age gradient in HRQoL, whereas adjustment for BMI alone resulted in only modest attenuation. In contrast, marginal effects of sex, race/ethnicity, education, and income changed little after adjustment for cardiometabolic conditions and/or BMI, with subgroup rank order and directional gradients preserved across models.

BMI and cardiometabolic conditions showed partially overlapping associations with HRQoL. Relative to models adjusting for cardiometabolic conditions only, additional adjustment for BMI modestly attenuated cardiometabolic condition-specific HRQoL decrements, most notably for obesity (−0.0305 to −0.0183), diabetes (−0.0427 to −0.0414) and high blood pressure (−0.0328 to −0.0316) in 2015, 2016, 2018, and 2022 MEPS (Table 2). Conversely, relative to models adjusting for BMI only, additional adjustment for cardiometabolic conditions attenuated the BMI-specific HRQoL decrements. The BMI coefficient indicated a change in HRQoL of −0.0022 per 1 kg/m² (attenuating to −0.0009 per 1 kg/m² after adjustment for cardiometabolic conditions). Despite this attenuation, both condition-specific and BMI-specific HRQoL decrements remained statistically significant in the fully adjusted model.

### 3.3 Temporal Trend on Cardiometabolic Condition-Specific HRQoL *Decrements*

Meta-regression indicated linear attenuation in condition-specific HRQoL decrements for diabetes, heart disease, and stroke from 2001 to 2022, whereas the high cholesterol-associated decrement became more negative (Figure 3; Online Supplement Table T). The diabetes-associated decrement attenuated from −0.0489 in 2001 to −0.0406 in 2022 (annual change +0.0007, P<0.0001). Similarly, the heart disease-associated decrement attenuated from −0.0591 in 2001 to −0.0493 in 2022 (annual change +0.0007, P<0.0001). Stroke-associated decrements also trended toward attenuation (−0.0839 in 2001 to −0.0577 in 2022), although the linear trend did not reach statistical significance (annual change +0.0005, P=0.0844). In contrast, the high cholesterol-associated decrement became more negative from −0.0135 in 2005 to −0.0234 in 2022 (annual change −0.0004, P=0.0185).

**Figure 3.**
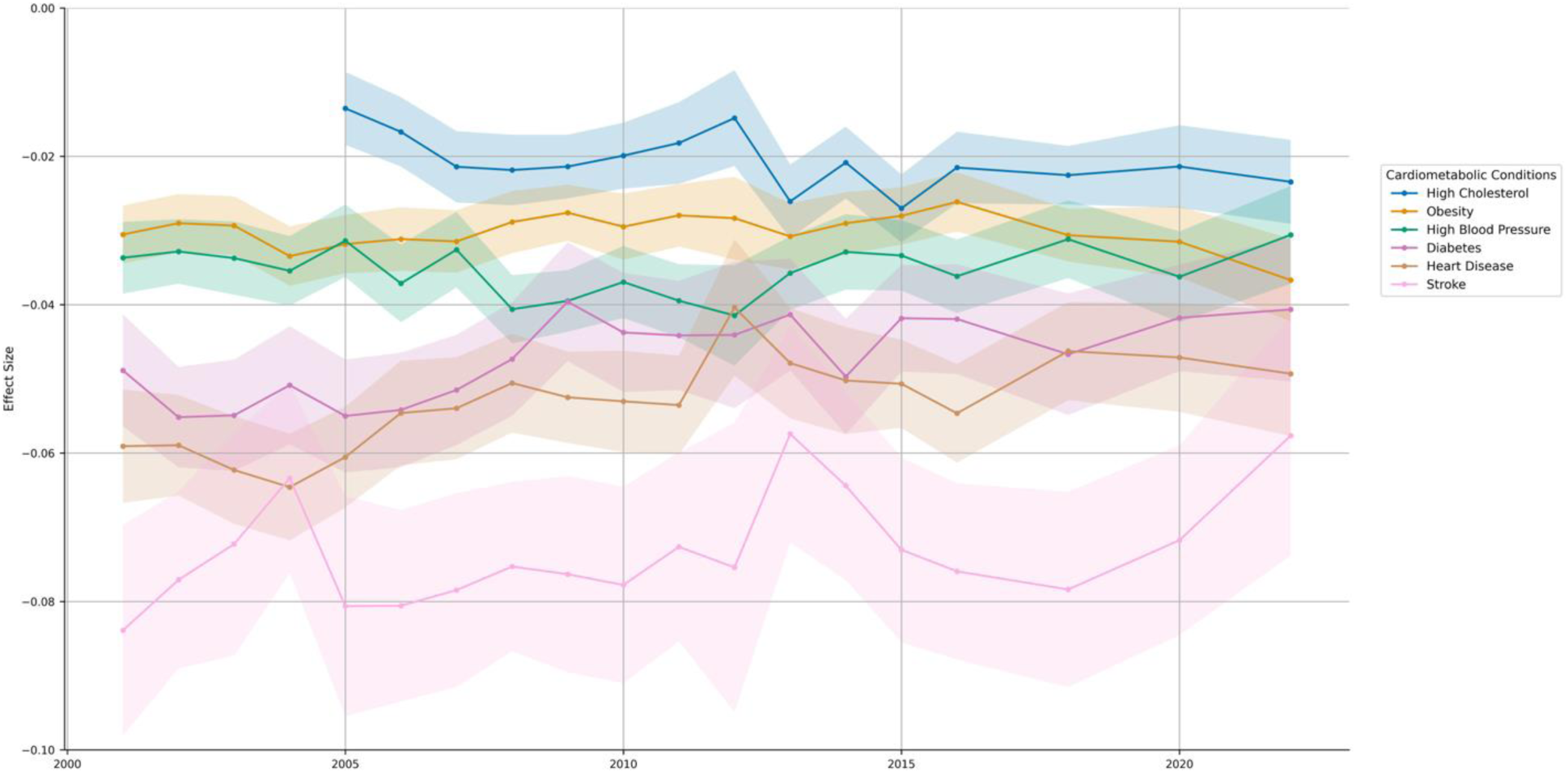
Cardiometabolic condition-associated HRQoL decrements over time, MEPS 2001-2022.

High blood pressure- and obesity-associated decrements exhibited nonlinear (quadratic) time patterns (Online Supplement Table U). High blood pressure showed the largest decrement around 2012 (−0.0337 in 2001, −0.0415 in 2012, −0.0306 in 2022), indicating smaller decrements both before and after 2012. Obesity showed the smallest decrement around 2012 (−0.0305 in 2001, −0.0283 in 2012) followed by worsening thereafter (−0.0367 in 2022).

## 4. Discussion

Using nationally representative MEPS data from 2001-2022 and an SF-12-to-EQ-5D mapping algorithm, we estimated associations between HRQoL and sociodemographic characteristics, BMI, and cardiometabolic conditions, assessed temporal changes in condition-specific HRQoL decrements, and generated nationally representative utility estimates for recent survey years. In adjusted models, accounting for cardiometabolic conditions substantially attenuated the age gradient in HRQoL, and improvements in condition-associated HRQoL over time were most evident among older adults. Adding BMI attenuated several condition-associated decrements. Over the study period, decrements associated with diabetes, heart disease, and stroke became less negative, whereas high cholesterol became more negative. Hypertension and obesity showed nonlinear patterns, with the most negative hypertension decrement around 2012 and more negative obesity decrements in later years.

Adjustment for cardiometabolic conditions substantially reduced age-group differences in HRQoL, suggesting that higher cardiometabolic burden at older ages contributes to lower HRQoL. A residual age gradient persisted, which may reflect other age-related conditions (eg, chronic kidney disease, cancer, cognitive impairment) that are independently associated with HRQoL.

Adjustment for BMI attenuated several cardiometabolic condition-associated HRQoL decrements, particularly for diabetes and high blood pressure, suggesting that part of the observed association between these diseases and HRQoL reflects shared risk factors related with obesity. This pattern is consistent with evidence that BMI may influence HRQoL both directly (eg, through increased depression or anxiety) and indirectly (eg, through raising the relative risk of cardiometabolic disease).^29,30^

These findings imply that when modeling interventions for cardiometabolic conditions that alter body weight (eg, lifestyle modification), utilities should consider (i) representing BMI as an explicit pathway to capture direct weight-related effects on HRQoL; (ii) avoiding double counting by assessing whether condition-specific utility estimates already adjust for BMI; and (iii) allowing BMI-HRQoL associations to vary by disease status when supported by empirical evidence (eg, different BMI-utility slopes among adults with vs without diabetes).^31^ More broadly, our results extend prior MEPS-based analyses of HRQoL, which examined associations with sociodemographic characteristics and clinical diseases but did not adjust for BMI despite its central role in cardiometabolic risk and population HRQoL.^23,32^

Diabetes-associated HRQoL decrements attenuated steadily from 2001 to 2022 (−0.0489 to −0.0406; annual change +0.0007, P<0.0001), indicating that the adjusted HRQoL difference between adults with and without diabetes became less negative over time. This pattern is consistent with national evidence of improving diabetes management and cardiovascular risk-factor control, including increased use of antihypertensive and statin therapy among eligible adults and higher attainment of guideline-based targets across the 2000s.^33^ Declines in major diabetes-related complications over decades (eg, acute myocardial infarction, stroke, amputation, and hyperglycemic crisis) provide a plausible pathway for improved population health status and functioning among adults living with diabetes.^34^ Our finding is consistent with prior MEPS-based evidence of improving physical health among adults with diabetes, supporting the plausibility of an upward shift in population HRQoL during this period.^35^

Heart disease-associated HRQoL decrements attenuated from 2001 to 2022 (−0.0591 to - 0.0493; annual change +0.0007, P<0.0001), indicating that the adjusted HRQoL difference between adults with and without heart disease became less negative over time. This pattern is consistent with substantial long-term declines in U.S. heart disease mortality through 2022 and a changing distribution of heart disease phenotypes, with fewer deaths from myocardial infarction and relatively greater contributions from other forms of heart disease, which may alter the functional status and symptom burden of people living with cardiovascular disease.^36,37^

Population surveillance data also suggest declines in angina symptom prevalence in U.S. adults over time, consistent with reductions in symptomatic coronary disease burden that would be expected to translate into higher health status.^38^

High blood pressure-associated HRQoL decrements reached their most negative values around 2012 (−0.0337 in 2001; −0.0415 in 2012; −0.0306 in 2022). This trajectory is consistent with national surveillance showing that hypertension control improved into the early 2010s but later declined, with more recent evidence suggesting partial recovery in control in subsequent years.^39^ This pattern may reflect changing symptom burden, complication risk, and functional limitations among adults living with hypertension. The 2017 ACC/AHA guideline’s emphasis on risk-based management and lower treatment thresholds may be consistent with improvements in health status among treated adults, although our analysis does not assess causal effects.^40^

Obesity-associated HRQoL decrements followed an inversed U-shaped temporal pattern, with the smallest decrement around 2012 (−0.0283) and more negative decrements thereafter (−0.0367 in 2022). This trajectory aligns with substantial increases in obesity prevalence and, notably, growth in severe obesity. From 1999-2000 to 2017-2018, the age-adjusted prevalence of obesity increased from 30.5% to 42.4%, and severe obesity nearly doubled from 4.7% to 9.2%.^41,42^ A larger share of class II/III obesity may contribute to larger utility losses in domains such as mobility and pain, consistent with the post-2012 pattern.

Overall, our condition-specific HRQoL estimates align with evidence from prior studies. A 2021 study reported mean EQ-5D utilities of 0.788 (SD, 0.233) for adults living with diabetes and 0.793 (SD, 0.223) for high blood pressure.^43^ Using 2000 MEPS data, a 2005 analysis estimated a 0.073 utility decrement for severe obesity versus normal weight.^46^ Earlier patient surveys from 2000-2001 found a mean EQ-5D of 0.80 among adults with diabetes,^44^ and a systematic review of stroke reported a pooled EQ-5D utility value of 0.68 (95% CI, 0.61−0.76).^45^ These comparisons support the face validity of our estimates.

Several limitations should be considered. First, we mapped EQ-5D utility values from SF-12 scores using a validated mapping algorithm rather than directly measuring them. Although this method is widely used and externally validated, prediction error and imperfect alignment with observed EQ-5D data may introduce bias. We evaluated face validity by comparing mapped EQ-5D trends with SF-12 trajectories and observed consistent patterns. Second, MEPS relies on self-reported BMI, which can lead to reporting bias. While correction methods exist for self-reported BMI, their use can introduce new biases and may not consistently improve population estimates.^47^ Third, MEPS defines “heart disease” broadly (including coronary heart disease, angina, myocardial infarction, and other heart disease), such that temporal changes in the estimated decrement may reflect evolving case mix and survivorship in addition to treatment effects; accordingly, we interpret heart disease trends as population-level associations rather than attributing changes to specific clinical pathways. Fourth, although we observed statistically significant attenuation in several disease-specific HRQoL decrements (eg, ≈+0.0007 HRQoL units per year for diabetes and heart disease), the annual change is small relative to a commonly cited threshold for clinically meaningful within-person change (∼0.03 utility units).^48^ To contextualize the potential relevance of these incremental shifts, we complemented trend estimates with a population-level illustration showing how modest per-person utility changes can accumulate into meaningful QALY differences when applied to highly prevalent, long-duration diseases (eg, diabetes). Finally, our analysis focused on single condition and did not fully incorporate multimorbidity or interactions between conditions. Future work should incorporate disease severity, clustering, and control, and should evaluate directly elicited utility values to strengthen evidence for policy and clinical decision-making.

## Data Availability

This study obtained data from 2001-2022 Medical Expenditure Panel Survey (MEPS) Full-Year Consolidated Public Use Files. The analysis code used in this study is openly available on GitHub.

https://github.com/yangda761/Cardiometabolic-Conditions-and-HRQoL

